# Exploring local government public health grant spending by health indicators, time and deprivation strata: an ecological study in England

**DOI:** 10.64898/2026.03.12.26348255

**Authors:** Estera Mendelsohn, Tom Prendergast, Talia Boshari, Caroline Fraser, Stefano Conti, Adam D M Briggs

**Affiliations:** The Health Foundation, London, UK; The Royal Free London NHS Foundation Trust, London, UK; London Borough of Camden, London, UK; Modelling and System Analytics, Data & Analytics, NHS England, London, UK; Faculty of Medicine, University of Southampton, UK

## Abstract

**Background:** The public health grant is used by upper-tier and unitary local authorities in England to fund public health services. Public health grant allocations have declined by 26% per person since 2015/16, with cuts being made without any adjustment based on population needs, resulting in absolute cuts often being greater for more deprived local authorities. This study seeks to investigate how these cuts have affected spending decisions across different areas of public health and how changes in spend relate to population health needs.

**Methods:** In this longitudinal ecological study, data on local government revenue expenditure and financing to 146 upper-tier local authorities in England were extracted from the Ministry of Housing, Communities, and Local Government for the years 2017/18, 2018/19 2019/20 and 2022/23. Demand for each function of the public health grant was proxied using a publicly available indicator of need.

Descriptive analyses explored changes to grant expenditure over time by function and IMD quintile. A compositional regression model was developed to account for the relatedness of spend data. The significance of associations between indicators of need and spend on functions of the grant was tested using MANOVA, producing Pillai’s Trace statistics as an indication of the effect size of each explanatory variable relative to others.

**Findings:** Public health grant spending reductions were widespread. More deprived local authorities often experienced deeper absolute cuts against a backdrop of greater need, with spend being protected across all IMD quintiles in only three areas: children’s 0 to 5 non-prescribed functions, health protection, and public mental health. In the multivariate regression, there was limited relationship between indicators of health need and patterns of grant spend between public health categories.

**Interpretation:** There is no clear relationship between potential indicators of need and expenditure of the public health grant in different reporting categories. Instead, spending decisions are being driven by other factors that may include historic spend, wider local priorities and financial pressures. These findings suggest a review of the public health grant formula to support local authority public health teams to more strategically apportion spend based on population health need.

- **What is already known on this topic**
  - Local authority public health teams in England receive a ring-fenced grant from central government which was originally based on an allocation formula that has not been updated since 2012/13.
  - The grant has been cut substantially over the past decade, often with larger absolute cuts for more deprived local authorities.
  - No previous study has investigated how public health teams allocate a diminishing grant across competing areas of public health need and how this may vary by deprivation.
- **What this study adds**
  - This study found limited evidence that indicators of health need have driven public health grant allocation in related spend categories, nor any differences by deprivation. Our analysis is the first to explore multiple indicators of need and to employ compositional regression to account for corelations between categories of grant spend.
- **How this study might affect research, practice and policy**
  - This study supports a review of the public health grant funding formula to better distribute the public health grant according to local population health need.

## INTRODUCTION

Local authority public health teams in England are funded through a ring-fenced grant (the ‘grant’) from the Department of Health and Social Care (DHSC).[1] The grant is used to fund a variety of services and programmes aimed at improving local population health and tackling inequalities. For example, NHS health checks, sexual health services, health visitors, and drug and alcohol services. Some of these functions are ‘prescribed’, meaning local authority public health teams must deliver them in a particular way, while the delivery of other ‘non-prescribed’ functions can vary based on local need and priorities.[1]

Spend from the grant is excellent value for money, estimated to provide an additional quality adjusted life year for just £3,800 in 2013/14 compared with £13,800 for services funded through the NHS.[2] Yet, between 2015/16 and 2025/26, the grant was cut by 26% per person in real-terms to £3.9bn, equivalent to around 2.0% of the NHS budget in England which also comes from DHSC.[3,4]

These real-terms cuts in the grant have led to significant reductions in spend across a range of funded services. For example, the amount spent by local public health teams on sexual health services fell in real terms between 2015/16 and 2025/26 by 32%, adult drug and alcohol services by 25% and adult obesity services by 20%.[4] At the same time, rates of preventable ill health from drugs, alcohol and obesity have all increased.[5]

Local authorities took over responsibility from NHS primary care trusts (PCTs) for commissioning and providing public health services in 2013/14, following the Health and Social Care Act 2012.[6]

To allocate the grant between the 152 upper-tier local authorities in England, DHSC’s Advisory Committee on Resource Allocation (ACRA) developed a formula to estimate relative need based on local criteria such as population size and age. Local authorities were allocated amounts in 2013/14 based on previous spend on public health services by local PCTs, with adjustments in 2013/14 and 2014/15 alongside overall increases of 5.5% and 5% respectively to ensure that gaps from the ACRA-calculated target were closed.[5,7,8]

The ACRA formula was updated again in 2016 to reflect routine data updates as well as provide stronger weightings for need related to premature mortality and substance use, but this new formula was never implemented. Instead, the grant was cut by 26% in real terms per person between 2015/16 and 2025/26, with reductions made as a percentage of the total grant without any consideration of the local health profile. This has meant that absolute cuts have disproportionately impacted more deprived local authorities where per-capita budgets were originally higher. For example, the per-capita budget of the most deprived local authority in England, Blackpool, is £43 less in 2025/26 than in 2015/16, compared to a £14 per capita cut in Wokingham over the same period.[4,5]

The cuts are likely to have contributed to a growing burden of ill health and premature death. For example, from 2009/10 to 2017/18, each 1% reduction in per capita public health spend was associated with a 0.15% increase in the prevalence of multimorbidity.[9] When looking at local government spend more broadly, each £100 reduction in per annum spend between 2013 and 2017 was found to be associated with an average decrease in life expectancy at birth of 1.3 months for men and 1.2 months for women, with disproportionate impacts on people living in more deprived areas.[10] Relationships have also been found between reductions in local authority spend and avoidable emergency admissions,[11] hospital admissions for nutritional anaemia,[12] and deaths or admissions from opioids.[13]

Few studies have investigated the relationship between specific areas of public health grant spend and related outcomes. For example, Liu *et al* examined the relationship between local government public health spend in 2013/14 and childhood obesity prevalence in 2016/17.[14] While higher levels of expenditure were not found to be associated with lower levels of obesity, small positive associations were identified between spend on physical activity and children’s public health programmes, and obesity levels in 4-5 year-olds three years later. Conversely, Roscoe *et al* found that while disinvestment from the public health grant in alcohol and drug treatment services were associated with a small but significant reduction in people accessing and completing treatment, there was no significant association with alcohol or drug related deaths, or alcohol-specific hospital admissions.[15]

These studies reflect not only the methodological difficulties with deriving causal relationships between public health grant spend to outcomes, but also the practical challenges faced by local government public health teams in managing declining budgets. Public health professionals need to consider local health needs and inequalities as well as the opportunity costs of investing more in one area of public health compared with another.

No study has systematically investigated how cuts to the value of the grant since 2015/16 have impacted public health spending decisions across different categories of expenditure and different public health functions, and how this varies between different local authorities and different public health needs. A better understanding of local authority public health spending decisions can help inform DHSC decisions about future grant allocations and whether the formula used for their distribution between local authorities should be reviewed.

This study aims to address this gap by quantifying:

1. how changes in spending on different public health reporting categories between 2017/18 and 2022/23 vary by local authority deprivation quintiles; and
2. how changes in spending on different public health reporting categories relate to changes in population health needs over time.

## METHODS

### Setting

We conducted a longitudinal ecological study of public health expenditure in unitary and upper tier local authorities in England in 2017/18, 2018/19 2019/20 and 2022/23 using publicly available data.

### Outcome measure

The outcome of interest was the amount of the public health grant spent across different public health reporting categories.[16] Data were collected from the Ministry of Housing, Communities and Local Government (MHCLG), which is publicly available and submitted by local authorities on a yearly basis.[17] The column ‘total expenditure’ was used rather than ‘net expenditure’ to separate any patterns in expenditure from an increase or a decrease in local authority income. Spending across the study period was adjusted to 2017 prices using the gross domestic product deflator.

### Explanatory measures

To understand whether public health funding allocations have been driven by population need, data on key health outcomes associated with categories of public health grant expenditure were collated. We then identified the most specific, publicly available indicators that could serve as proxies for demand for each category. For example, child obesity prevalence was used as a proxy for need for spend within the reporting category, ‘obesity – children.’ In cases where a clear indicator of need was not available, we used the relevant population numbers as a proxy for service demand. For example, the number of children aged 0 to 5 in each local authority was used to estimate spend within the ‘children’s 0 to 5 services’ category. To account for a delay between data collection on indicators of need and any resulting changes to spend, the exposures (indicators of need) were linked to the outcome (changes to public health grant spend in each category) by a two-year time lag, as well as a one- and three-year lag in sensitivity analyses. Table 1 summarises the reporting categories of the public health grant and their corresponding indicator of need.

**Table 1.**
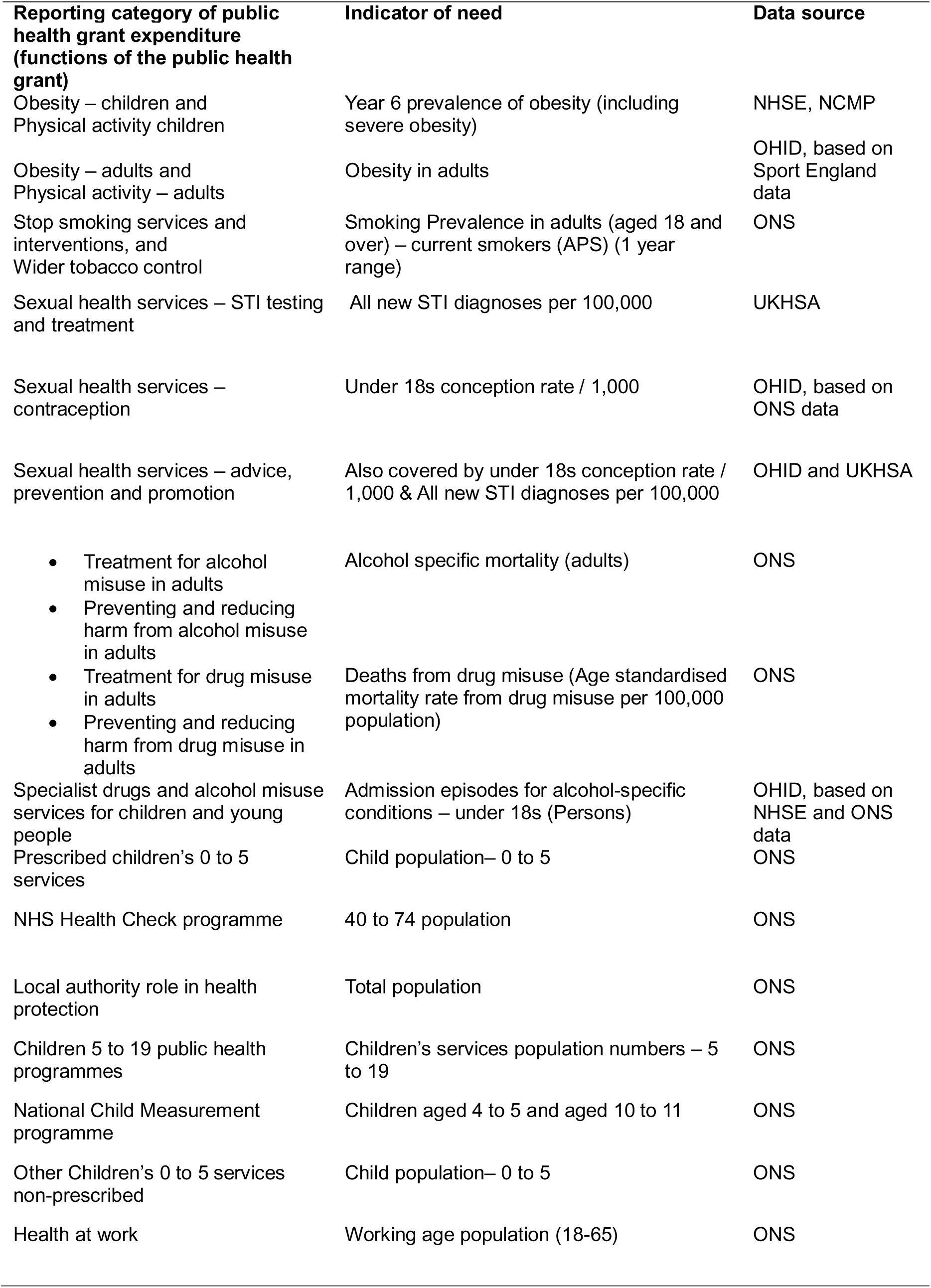
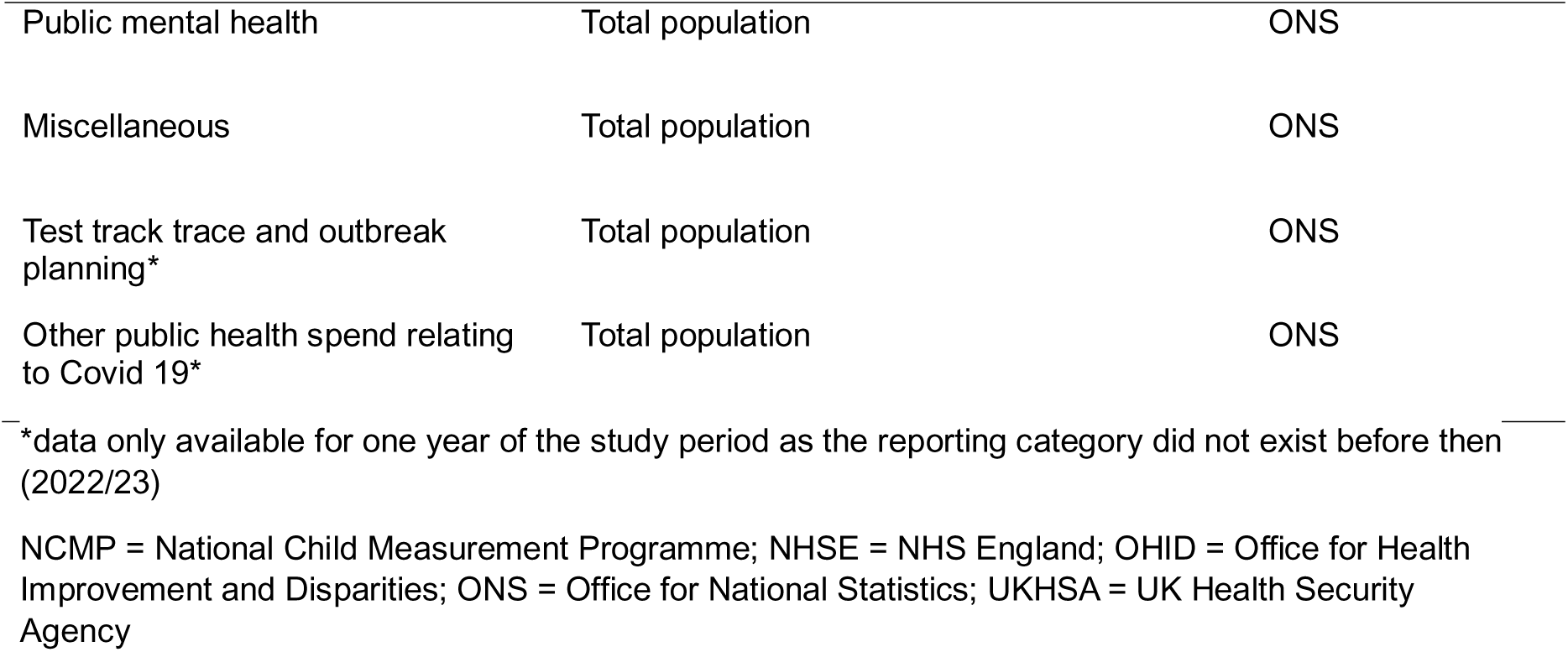
Functions of the public health grant and the associated indicator of need used in the analysis. Reporting categories of the grant which served a similar purpose were grouped together for analysis.

### Missing data

Missing data was minimal across local authority expenditure reports (see supplemental appendix 1). However, spending £0 for a particular function was more common in the dataset. It was not possible to explain whether this was due to a true value, a lack of data submitted, or an error. Most local authorities (n=135) contained at least a single function with £0 spend across the study period. The median number of categories with £0 spend across the local authorities affected was three in 2016 and two in the remaining years of the study period.

Some key indicators were only available from 2015 onwards, specifically obesity prevalence in adults, and admission episodes for alcohol-specific conditions. As full sets of data were required for our inferential analysis (compositional regression), this meant that spending years 2015/16 and 2016/17 (with indicators of need stemming from 2013 and 2014) were excluded from the analysis. The final period of study for the regression analysis was therefore from 2017/18-2019/20 and 2022/23. Spending data for the financial years 2020/21 and 2021/22 were excluded from the analysis due to Covid-19-related adjustments to the public health grant, meaning spending decisions were ungeneralisable.

Where data points were missing in the years selected for the regression, values were imputed using k-nearest neighbour estimation.[18] Twelve local authority codes were removed because they corresponded to local authorities which were dissolved, merged, newly created during the observed period, or did not submit spending data in one or more of the selected years (see supplemental appendix 1).

Prevalence of Year 6 obesity was missing for the year 2020 (acting as an indicator of need for spending year 2022/23) in 121 local authorities, likely due to school disruption during the COVID-19 pandemic. As elsewhere, these missing 2020 values were imputed using k-nearest neighbour estimation. For more information on missing data, please see supplemental appendix 1.

### Descriptive statistics

We explored changes to public health grant expenditure in both absolute and relative terms, which is recommended when considering health inequalities.[19] We initially mapped the absolute changes in median spend (as £ per capita) across functions of the public health grant during the study period by IMD quintiles using descriptive statistics. We used the midyear population estimate for each local authority, obtained from the Office for National Statistics (ONS) to estimate the relevant population. The denominators used for per capita calculations for each grant function are set out in supplemental appendix 2. Similarly, we explored changes to the explanatory variables, key indicators of demand, for each function by IMD quintile (Figure 2, supplemental appendix 3).

### Compositional data model

As this study aimed to understand changes in the allocation of the grant across 21 different functions, the outcome of interest is considered multivariate (i.e., the analysis involves two or more variables at the same time). Moreover, because the public health grant is distributed across services that together form a whole (100% of the grant), the data are also *compositional* in nature. Compositional data differ from traditional multivariate datasets in that an increase in one category must be accompanied by a corresponding decrease in another, which can lead to spurious correlations. As all functions are allocated funds from a fixed grant, the proportion spent in one area is intrinsically dependent on the choices made about spending in other areas. As a result, analysing local authorities’ expenditure on isolated functions of the grant has little meaning without understanding the relative effects on other services. Compositional data must therefore be analysed using methods that account for their structure. As best practice, this is achieved by expressing compositions as log-ratio coordinates, which can then be used in traditional statistical models.[20]

Using the R package ‘compositions,’ isometric log-ratio (ilr) transformations were applied (using the *ilr* function) to the data to produce a set of ilr coordinates representing the percentage of the public health grant allocated to each reporting category.[21]

The unit of analysis was the local authority. To account for the repeated measures across time, year of spending was included as a fixed effect. Lastly, several covariates, hypothesised to confound the association between the amount of the grant allocated to a given function and health outcomes, were selected *a priori*. Area deprivation is a notable confounder because local authorities with higher levels of deprivation often face a higher prevalence of risk factors for adverse health which may lead them to allocate resources differently compared to areas of lower deprivation. Deprivation was based on the 2015 Index of Multiple Deprivation (IMD) scores, the most up to date IMD index at the time that most spending decisions were made.[22] Local authorities were ordered by the rank of proportion of LSOAs in the most deprived 10% nationally and divided into quintiles to create a categorical variable included in the model as a covariate. The final model was adjusted for year of expenditure, IMD quintile, urbanisation category of local authority, under 18s conception rate per 100,000, year 6 prevalence of obesity (including severe obesity), alcohol specific mortality, all new STI diagnoses per 100,000 people, smoking prevalence in adults, obesity prevalence in adults, deaths from drug misuse, admission episodes for alcohol specific conditions in under 18 year olds, total adults, 0 to 5 year olds, 40 to 74 year olds, 5 to 19 year olds, 4 to 5 and 10 to 11 year olds, and over 65 year olds population numbers (with all time-varying covariates linked by a lag of two years).

The significance of associations between indicators of need and the composition of public health grant spend was tested using multivariate analysis of variance (MANOVA – for more information on the application of MANOVA to compositional data.[23] For each explanatory variable *i*, significance was gauged against the null hypothesis that the association remained significant when jointly controlling for all the other *i*-1 variables included in the model, with each variable sequentially tested against all others. The test statistic used in this context is Pillai’s Trace, which also acts as an indicator of the rough effect size of each explanatory variable relative to others. The proportion of variance in composition of the public health grant explained by the model as a whole was measured using adjusted-R^2^ (for more detail see Gerald Van Fen Boogaart and Tolosana-Delgado, chapter 5).[23]

Spend related to COVID-19 was not considered as part of our regression analysis. In years where COVID-19 spending was present in local authority spending records, it was removed and the proportion made up by each other category of spending was re-calculated based on the total spend net of COVID-19 spending.

To further assess whether the composition of public health grant spending was influenced by need, we performed analyses focussing on the sensitivity of thematically unified portions of public health spending to our selected indicators of need. For these analyses, items of spending in the grant were divided into five thematic sub-groups as follows:

- *Children and young people-related spend:* encompassing spend on public health programmes for children 5-19, prescribed and non-prescribed functions for children aged 0-5, and prescribed functions relating to the national child measurement programme.
- *Substance misuse and tobacco control spend*: spend on substance misuse prevention and treatment for alcohol and drugs for adults, substance misuse prevention and treatment for alcohol and drugs for children and young people, smoking and tobacco - stop smoking services and interventions and smoking and tobacco - wider tobacco control.
- *Sexual health spend*: Spend on prescribed STI testing and treatment and contraception services, and non-prescribed sexual health advice.
- *Obesity-related spend*: Spend on obesity and physical activity for children and adults.
- *Other spending*: Spend on health at work, NHS health check (prescribed functions), local authority role in health protection (prescribed functions), miscellaneous public health services, public health advice (prescribed functions), and public mental health.

Additional compositional regressions were run assessing the influence of the indicators of need on the balance of each of these sub-groups of spend individually compared to everything else contained in the grant.

### Sensitivity analyses

Sensitivity analyses were performed on the selected time-lag between indicators of need and year of spend (set to two years for the core analysis). The lag was both extended to three years and shortened to one year. This was to check whether any associations found between indicators of need and spending composition were still present or differed with the timing of the indicator relative to the time of spend, adding to our understanding of spending decisions’ responsiveness to need. Sensitivity analyses can be found in supplemental appendix 3.

## RESULTS

### Descriptive results

Public health grant expenditure varied significantly by function, with average local authority annual spend dominated by four categories (Figure 1). The biggest category of spend was children’s 0-5 services which includes health visitors and where local authorities collectively spent, on average, £684m each year over the four-year study period. Conversely, functions such as ‘health at work,’ the ‘national child measurement programme prescribed functions,’ and health protection cumulatively accounted for less than 2% of overall spend.

**Figure 1.**
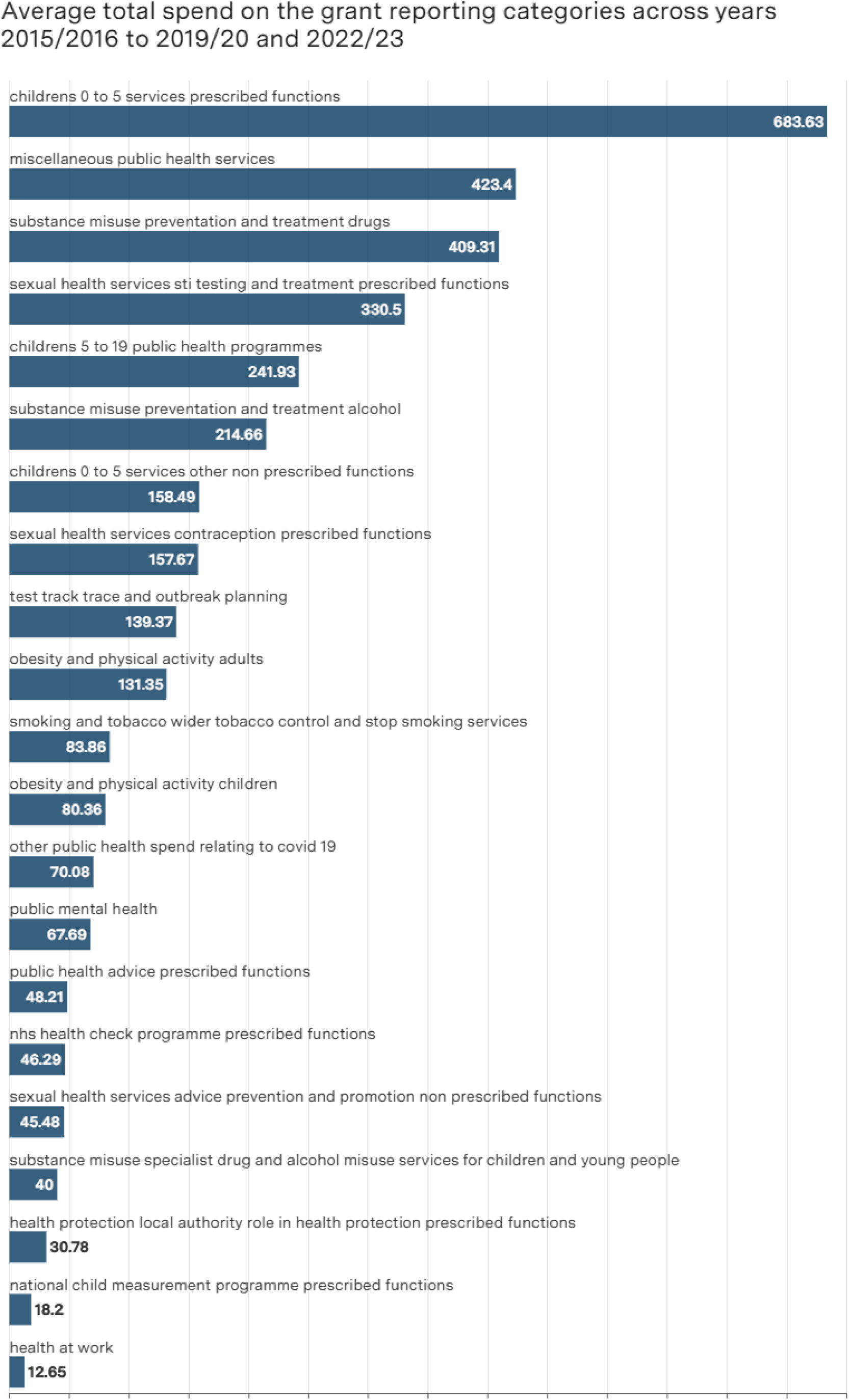
Average annual spend per function of the public health grant. Average annual spend was calculated by summing the total spend across local authorities for a given function and divided by the number of years that the function was reported. COVID-19 related functions were only reported in 2022/23. All other functions were reported across the 4-year study period.

Since 2017, per capita grant expenditure declined across most categories. Spend was maintained in only three categories across all IMD quintiles: Children’s 0 to 5 non-prescribed functions, health protection, and public mental health. Spend was also maintained in some, but not all, IMD quintiles in several other categories, such as the national child measurement programme, childhood obesity and physical activity categories, and drug and alcohol misuse services for children and young people.

Areas with higher deprivation continued to spend more per person than areas with lower deprivation across all categories, with the exception of NHS Health Checks. In several categories, per capita spending increased in the least deprived IMD quintile, but declined in the most deprived quintile, including health at work, adult obesity and physical activity, sexual health services (advice, prevention and promotion, non-prescribed), and alcohol misuse prevention and treatment in young people.

In some categories, trends in population need mirrored trends in expenditure for grant categories (Figure 2). For example, reductions in spending on STI services and contraception services, coincided with declines in STI incidence and under 18s conceptions, while decreased spending on smoking services was seen alongside a decline in smoking prevalence. However, deaths from alcohol and substance misuse, and obesity prevalence increased over the period of interest against a backdrop of reduced or stagnant spend on obesity services and alcohol and substance use services.

**Figure 2:**
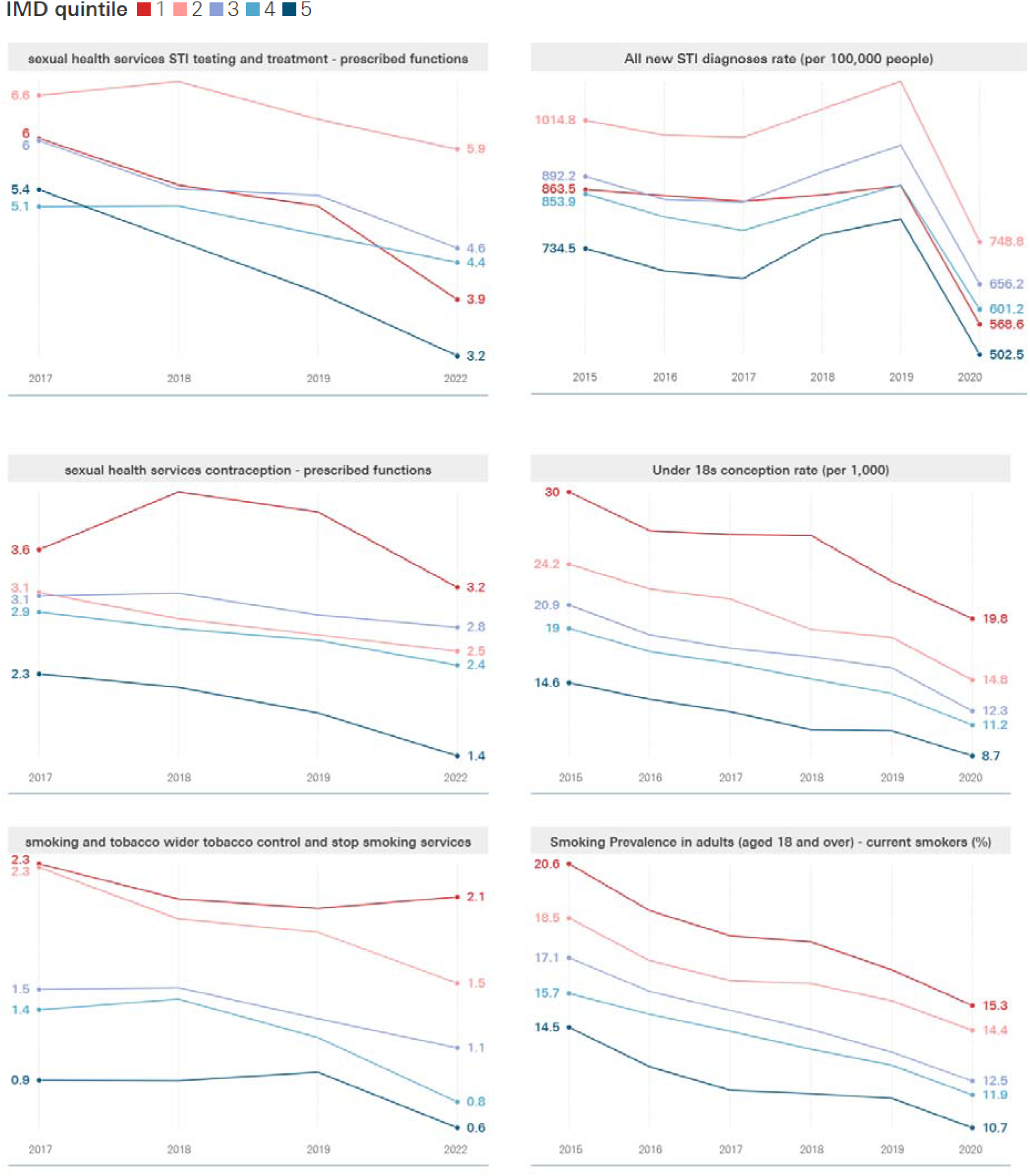
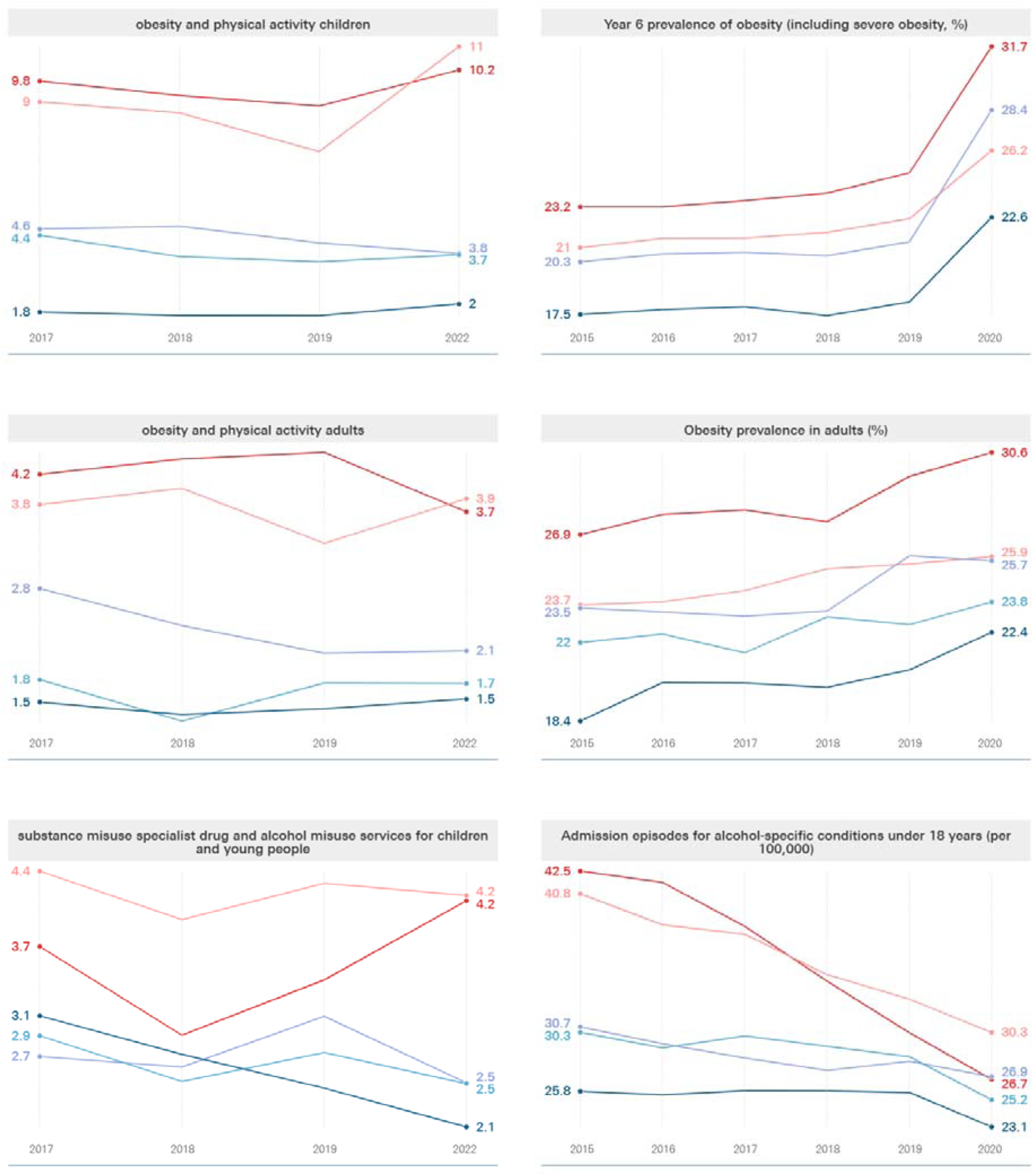
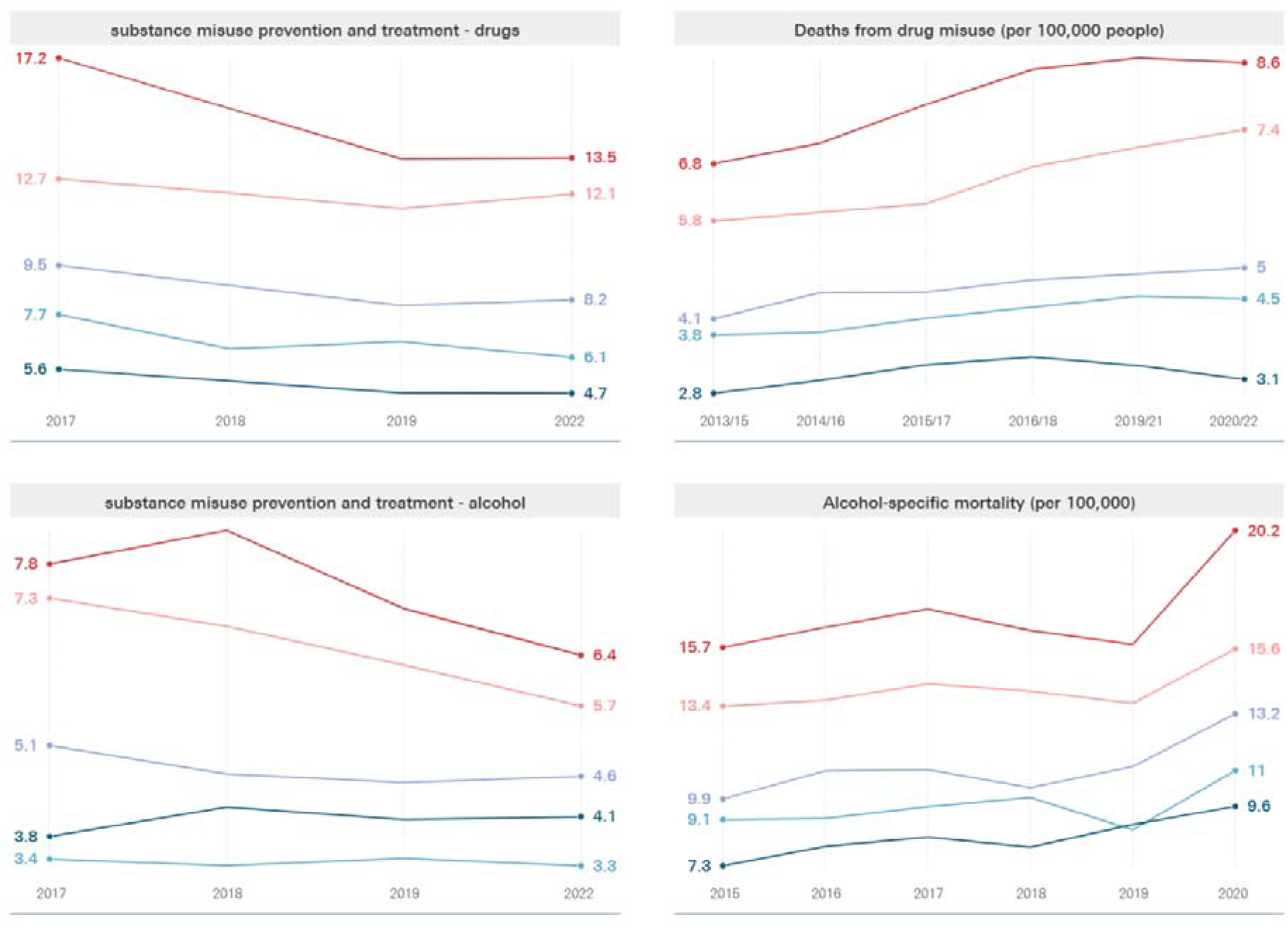
Panel showing median spend per capita per function of the public health grant, stratified by the IMD quintile of local authorities (left) compared to values of indicators of need (right). IMD quintile 1 (red) indicates highest deprivation while quintile 5 (blue) least. Data for financial years 2020/2021 and 2020/2021 is omitted. Denominators used to calculate per capita spend are reported in supplemental appendix 2 Table A2.1.

### Regression results

Most indicators of need were found to be significantly associated with how the grant was allocated between reporting categories (Table 2). The only exceptions to this were the measures of admission episodes for alcohol specific conditions for those under the age of 18 and the proportion of the population between the ages of 4 and 5 or 10 and 11 (used as an indicator of need for spend on the National Child Measurement Programme), for which no evidence of association was found. The largest effect sizes (measures as Pillai’s Trace test statistic) were observed for urbanisation category and IMD quintile by a considerable margin, suggesting that these factors more strongly influence the composition of public health spending.

**Table 2.** MANOVA summary assessing the association between two-year-lagged indicators of need and the composition of the public health grant.

| Variable | Pillai's Trace | P-value |
| --- | --- | --- |
| Urbanisation category | 0.43 | <0.01*** |
| Year | 1.03 | <0.01*** |
| IMD quintile | 0.40 | <0.01*** |
| Under 18s conception rate per 1,000 | 0.06 | 0.01*** |
| Year 6 prevalence of obesity | 0.12 | <0.01*** |
| Alcohol specific mortality | 0.06 | 0.04*** |
| STI diagnoses rate per 100,000 | 0.07 | <0.01*** |
| Smoking prevalence in adults | 0.07 | <0.01*** |
| Obesity prevalence in adults | 0.06 | 0.03*** |
| Deaths from drug misuse | 0.8 | <0.01*** |
| Admissions for alcohol specific conditions (<18 yrs) | 0.03 | 0.45 |
| Adult population percentage | 0.12 | <0.01*** |
| Age 0-5 population percentage | 0.11 | <0.01*** |
| Working age population percentage | 0.09 | <0.01*** |
| Age 40-74 population percentage | 0.15 | <0.01*** |
| Age 5-19 population percentage | 0.13 | <0.01*** |
| Age 4-5 or 10-11 population percentage | 0.04 | 0.15 |
| Age over 65 population percentage | 0.17 | <0.01*** |

Note: Pillai's trace statistics give an indication of effect size, but they do not directly quantify the change in our outcome associated with a unit change in an exposure variable, and should not be interpreted as such.
\*\*\* indicates significance to the 95% level.

Although most of these indicators of need were found to have a significant association with the composition of the public health grant, they only explain a small amount of the variation in public health grant composition between local authorities (23% of total variation explained; R^2^ = 0.23). This implies that most variation in spending patterns is likely to be driven by other factors rather than the measures of need explored in this study.

There remains the possibility that certain categories of grant spend are more sensitive to changes in need than others, or more responsive to some indicators of need over others. This was assessed through additional regression analyses which categorised functions into thematic sub-groups. For each of these, urbanisation category, IMD quintile and the proportion of the population over the age of 65 were found to have a significant influence on the composition of the grant at the sub-group level. In all sub-groups, the largest effect size (gauged from Pillai’s trace) came from urbanisation category and IMD quintile – reflecting what was observed from function-level regression results. The size of these effects did vary between sub-groups, however, with urbanisation having a larger effect on substance abuse, sexual health, and other spending (Pillai’s Trace of 0.12, 0.11, and 0.11 respectively) than it did on obesity or child-related spend (0.05 and 0.07 respectively). Additional variables were also found to have a significant association with the proportion of each sub-group of spend versus the rest of the grant, but with no clear pattern of which categories most drive spending decisions (see supplemental appendix 5).

Sensitivity analyses were performed where the lag between indicators of need and the composition of the public health grant was altered (lengthened from two to three years, and shortened to one year). Few differences were observable between the results of the primary analysis and these sensitivity analyses (see supplemental appendix 4).

## DISCUSSION

Following its transfer to local authorities, public health spend has been dominated by three macro functions of the grant: children’s 0-5 services prescribed functions, alcohol and substance use services, and sexual health services (STI testing and treatment), as well as spend that falls within the broad ‘miscellaneous’ public health services category. Per capita spend was generally higher among more deprived local authorities compared with less deprived local authorities at the start and end of the study period. However, fluctuations in the intervening years do not present a clear or consistent trend across different functions of the grant or by IMD quintile.

The regression results present a similarly mixed picture. The Pillai’s Trace statistics of two variables – IMD and urbanisation – are demonstrably larger than others, signalling the relative strength of these upstream determinants of public health need in affecting public health grant allocations over indicators related to specific spending categories such as obesity or smoking rates. However, the strength of this association should be interpreted with caution as all variables collectively account for just 23% of overall variation in expenditure. Furthermore, spend on thematic sub-groups of public health functions is related to a range of indicators of need, some of which are not directly related to the thematic sub-group. For example, sexual health spend is not influenced by the rate of new STI diagnoses, and obesity spend is related to obesity rates in year 6 children but not among adults. Finally, changes to public health grant spend by category did not meaningfully differ when linked to indicators of need by a one-, two- or three-year time lag, further implying a weak correlation between spending decisions and measures of public health need.

Together, these findings show no clear relationship between potential indicators of need and expenditure of the public health grant against different reporting categories. Instead, cuts to the grant over the past decade have likely been managed based on other factors that may include historic spend, alternative local priorities and broader financial pressures. This lack of relationship between need and spending decisions adds weight to the argument that the public health grant funding allocation formula for local authorities should be reviewed to better meet local needs. One such opportunity would be the current UK government consultation on the allocation of central government funding for local authorities in England, including introducing new assessments of need and potential for local revenue raising.[24] While the proposals involve consolidating several grants for public health services, it appears that the approach to distributing these funds will not change from previous years.

Previous studies have explored the impact of reduced public health grant spend on specific health outcomes, but few have aimed to understand whether spending decisions are influenced by, or related to, pre-existing information on health risk factors. Studies that have analysed the public health grant at the level of individual functions have observed an inconsistent relationship between expenditure on a particular function and indicators of need.[14,15] However, these studies analysed categories of grant spend in isolation, rather than treating them as part of a whole with methods that can handle compositional data. Furthermore, Liu *et al* and Roscoe *et al* looked prospectively at whether changes in function-level spend affected health outcomes, whereas our study considers the reverse: how health outcomes may be driving spend decisions.[14,15]

The main strengths of our study lie in the strong conceptual justification of the research question, and the methods used to explore it. Compositional regression is an emerging technique in public health. While it has recently gained attention in areas such as physical activity research and movement behaviours,[25,26] we are the first to demonstrate its utility in exploring healthcare spending as a compositional outcome. Amid increasing budgetary pressures in many countries, this method can offer insights into whether resources are distributed where they are most needed. Additional strengths of our methodology include the broad range of variables considered as possible drivers of spend, as well as the completeness and reliability of the datasets, which were sourced from government publications.

There remain several limitations to our study. Firstly, there is potential for unobserved confounding between local authorities. For example, local authority spend external to the public health grant in areas such as housing, infrastructure, social care and education (critical wider determinants of health) was not accounted for in our research, yet spend in these areas will impact local population health and may influence grant allocation decisions.

Secondly, coefficients from compositional regression do not directly correspond to real-world effect sizes, meaning they cannot be used to quantify absolute changes in category-level spending. While our analysis identifies variables which influence the overall composition of public health grant expenditure, it cannot identify which specific functions of the grant are directly affected by each indicator of need or the direction of the influence that different indicators exert on the grant’s composition. For example, although our results show that urbanisation affects how public health grant funding is allocated locally between services, we cannot tell which aspects of the grant receive more or less funding due to differences in this variable, or how large we would expect these differences to be. Therefore, while we identify determinants of the grant’s composition, their precise relationship to different categories of the grant requires further investigation. This reflects broader limitations in the compositional regression literature, which remains methodologically fragmented and lacks widely adopted software packages to support applied use, particularly for compositional dependent variables with more than three parts.[26,27] Future research could explore more novel variations of this method which may support clearer interpretation.[27–29]

Finally, it is possible that erroneous data entries remained in the source dataset despite steps taken to ensure local authorities with inconsistencies were removed from the analysis. Reported spending of £0 by local authorities was taken to be a true value, rather than missing data. While the data comprehensively covers England and our findings may be relevant to other high-income countries where preventative public health is exposed to fiscal constraint, their generalisability to international settings is likely limited by differences in health system financing structures.

## CONCLUSIONS

Our study suggests that indicators of population health need are weakly related to public health grant spend across different public health categories. Instead, between 2017/18 and 2022/23, local authorities managed spending decisions alongside cuts in the public health grant based on other factors which may include historical spend and alternative local funding priorities and pressures. Further qualitative research should explore the different potential factors that influence local authority public health teams’ spending decisions. This would support the growing body of evidence justifying a review of the public health grant formula to better align grant allocations with the different needs of councils in England. This could better support local authority public health teams to more effectively deliver services that meet local population needs and avoid exacerbating persistent health inequalities.

## Footnotes

### Contributions

**EM:** Formal analysis; Data curation; Methodology; Validation; Visualisation, Writing (Original Draft Preparation), Writing (Reviewing & Editing). **TP:** Formal analysis; Methodology; Validation; Writing (Original Draft Preparation), Writing (Reviewing & Editing). **TB:** Writing (Original Draft Preparation), Writing (Reviewing & Editing). **CF:** Data curation; Writing (Reviewing & Editing). **SC**: Methodology. **AB:** Conceptualization; Project Administration; Supervision; Writing (Original Draft Preparation), Writing (Reviewing & Editing). AB accepted full responsibility for the finished work and/or the conduct of the study, had access to the data and controlled the decision to publish.

## Acknowledgements

We are grateful to Hugh Alderwick, Director of Policy and Research, The Health Foundation, for reviewing and editing this paper.

## Data availability statement

Data were available to the public before the initiation of this study. Data can be downloaded from Ministry of Housing, Communities & Local Government, Office for national statistics and the Department of Health and Social Cre (Fingertips).

## Ethics statement

Patient consent for publication: **Not applicable**.

