## Supplementary Material for "Exploring local government public health grant spending by health indicators, time and deprivation strata: an ecological study in England"

**Appendix 1: Missing data**

The tables below show categories for which data were missing in some cases.

Table A1.1 Summary of missing data on public health grant spend.

| Issue | Categories affected | Local authorities affected |
| --- | --- | --- |
| No data for whole local authority in 2015 | All | North Yorkshire |
| No data for whole local authority in 2022 | All | Birmingham  Slough  Thurrock  Torbay |
| No data for whole local authority past 2015 | All | Bournemouth,  Christchurch & Poole,  Buckinghamshire Council,  North Yorkshire |
| Data only available in 2022 | All | Dorset  North Northamptonshire  West Northamptonshire |
| Only 2022 data available | - test track trace and outbreak planning - other public health spend relating to COVID-19 | All |
| No 2015 data available | - Public mental health - health at work | All |

Table A1.2 Summary of missing data on indicators of need, after excluding merged or dissolved local authorities.

|  | **Number of missing values** | | | |
| --- | --- | --- | --- | --- |
| **Indicator** | 2015 | 2016 | 2017 | 2020 |
| Under 18s conception rate per 1,000 | 4 | 4 | 4 | 1 |
| Year 6 prevalence of obesity | 4 | 4 | 4 | 131 |
| Alcohol specific mortality | 9 | 13 | 11 | 4 |
| STI diagnoses rate per 100,000 | 3 | 3 | 3 | 0 |
| Smoking prevalence in adults | 4 | 4 | 4 | 1 |
| Obesity prevalence in adults | 3 | 3 | 3 | 0 |
| Deaths from drug misuse | 11 | 11 | 11 | 6 |
| Admissions for alcohol specific conditions (<18 yrs) | 5 | 7 | 8 | 3 |

Prevalence of Year 6 obesity was frequently missing for the year 2020, likely due to school disruption during the COVID-19 pandemic. In all cases, missing data were imputed using k-nearest neighbour estimation with k = 2. A k of 2 was chosen as in most cases only a single year of data was missing in a series, meaning that almost all missing values would have 2 contiguous and likely similar values in its relevant time series.

Some local authorities were dissolved, created, or merged during the observed period, or did not submit spending data in one or more of the selected years. These were excluded from the analysis. In many cases these overlap with the authorities for which data was missing in one or more years, in Table A1.1. The excluded local authority codes were:

- E06000053
- E06000058
- E06000060
- E10000022
- E06000062
- E06000061
- E06000059
- E06000027
- E06000028
- E06000029
- E06000057
- E08000037

**Appendix 2. Per capita spend calculations**

Table A2.1. Denominators used to calculate per capita spend across functions of the public health grant.

| **Category** | **Denominators** |
| --- | --- |
| Children 5 19 public health programmes | Population of children aged 5-19 |
| Health at work | Total population aged 16 and over |
| Health protection local authority role in health protection prescribed functions | Total population |
| Miscellaneous public health services | Total population |
| Miscellaneous public health services children's 0 5 services other non-prescribed functions | Population of children aged 0-5 |
| Miscellaneous public health services children’s 0-5 services prescribed functions | Population of children aged 0-5 |
| National child measurement programme prescribed functions | Population of children aged 4-5yrs and 10-11 years |
| NHS health check programme prescribed functions | Population of adults aged 40-74 |
| Obesity adults | Total adult population |
| Obesity children | Total under 18 population |
| Other public health spend relating to COVID-19 | Total adult population |
| Physical activity adults | Total adult population |
| Physical activity children | Total under 18 population |
| Public health advice prescribed functions | Total population |
| Public mental health | Total population |
| Sexual health services advice prevention and promotion non prescribed functions | Total population |
| Sexual health services contraception prescribed functions | Total population |
| Sexual health services STI testing and treatment prescribed functions | Total population |
| Smoking and tobacco stop smoking services and interventions | Total population |
| Smoking and tobacco wider tobacco control | Total population |
| Substance misuse preventing and reducing harm from alcohol misuse in adults | Total adult population |
| Substance misuse preventing and reducing harm from drug misuse in adults | Total adult population |
| Substance misuse specialist drug and alcohol misuse services for children and young people | Total under 18 population |
| Substance misuse treatment for alcohol misuse in adults | Total adult population |
| Substance misuse treatment for drug misuse in adults | Total adult population |
| Test track trace and outbreak planning | Total population |

**Appendix 3.**

*Panel showing median spend per capita per function of the public health grant, stratified by the IMD quintile of local authorities (left) compared to values of indicators of need (right). IMD quintile 1 (red) indicates highest deprivation while quintile 5 (blue) least. Data for financial years 2020/2021 and 2020/2021 is omitted. Denominators used to calculate per capita spend are reported in supplemental appendix 2 Table A2.1.*

| 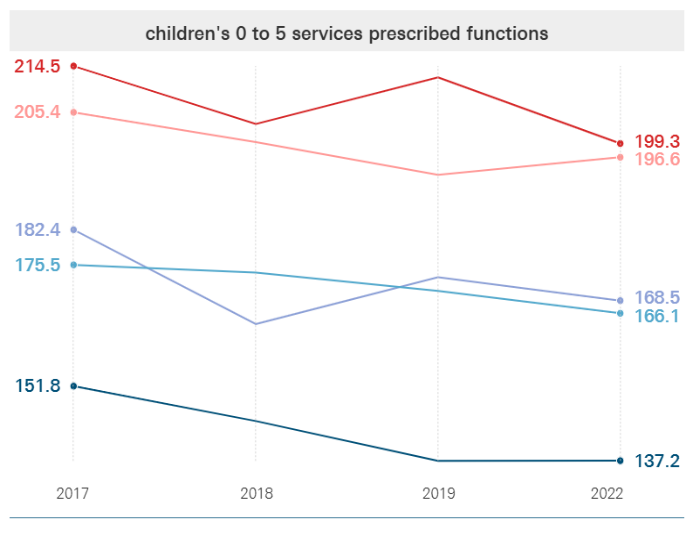 | 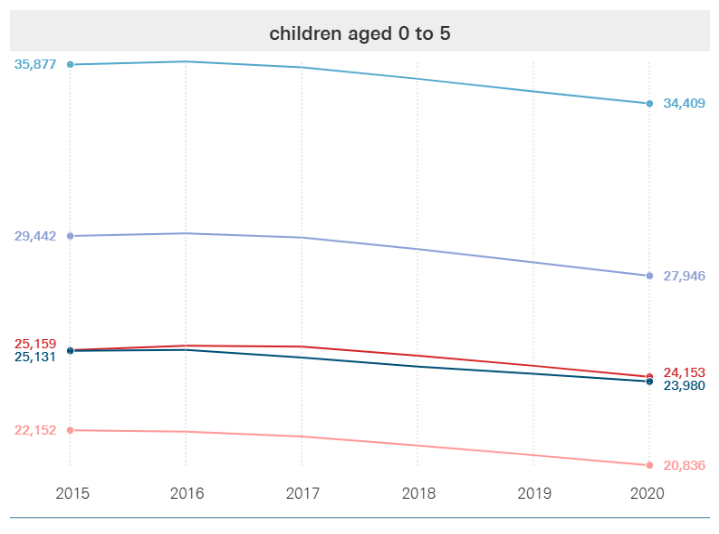 |
| --- | --- |
| 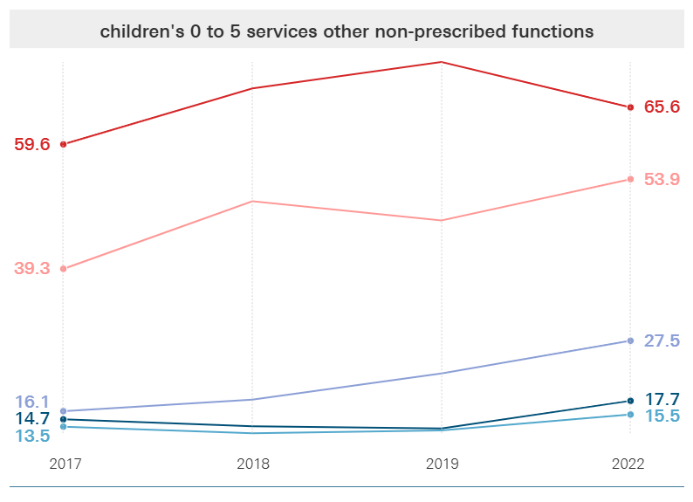 | 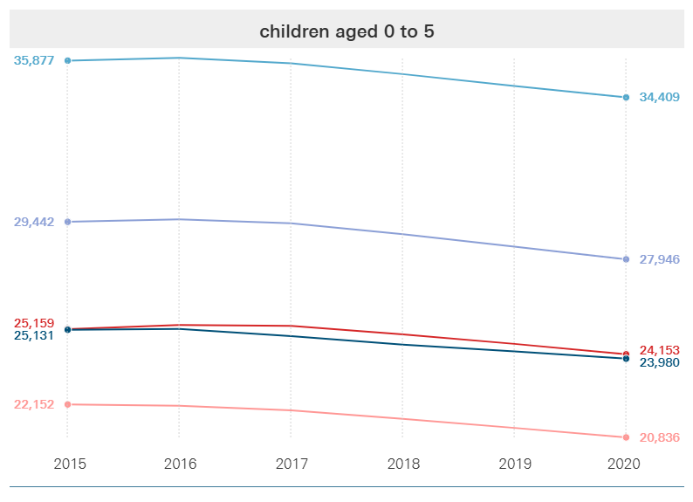 |
| 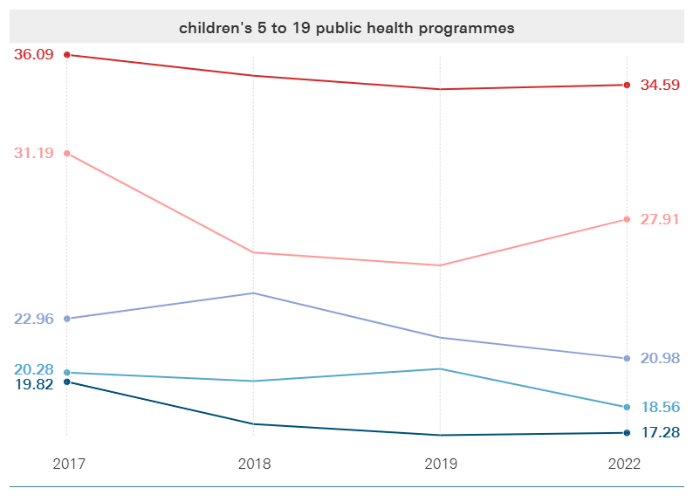 | 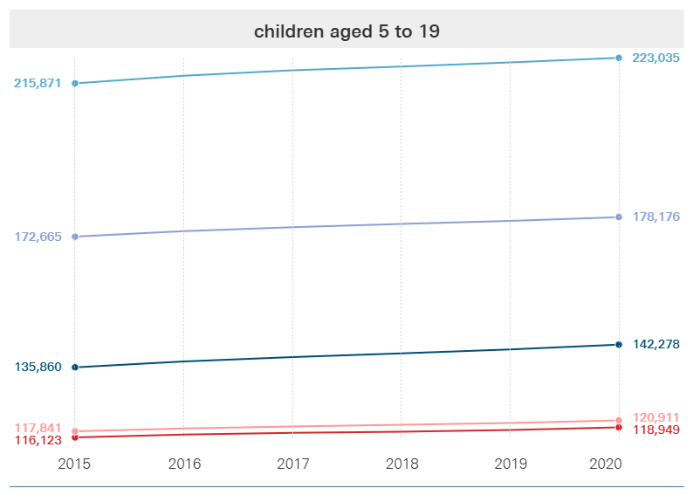 |
| 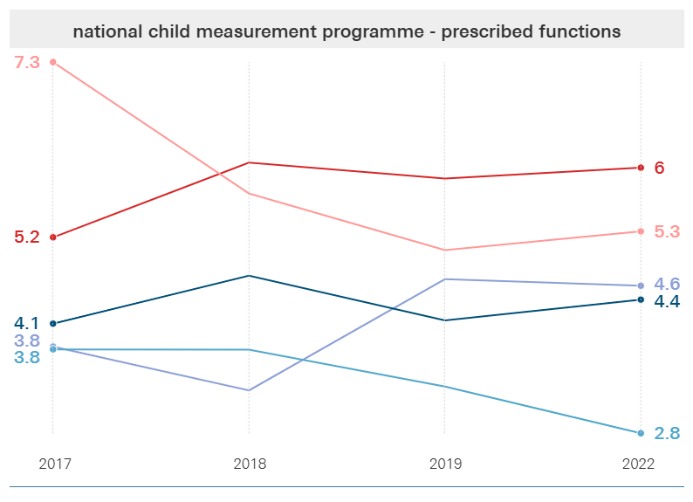 | 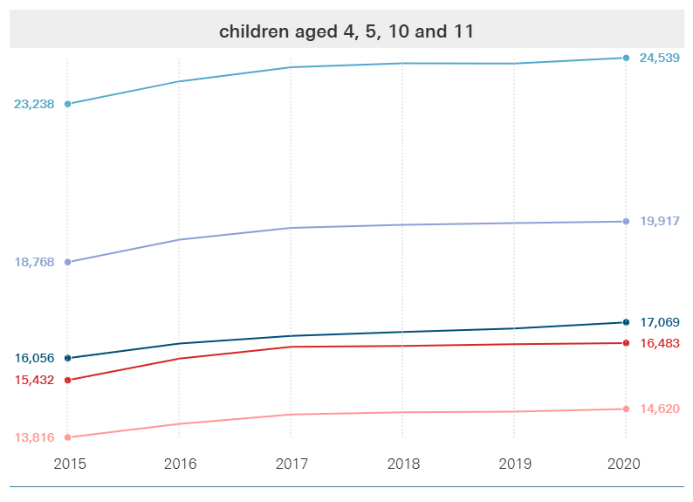 |
| 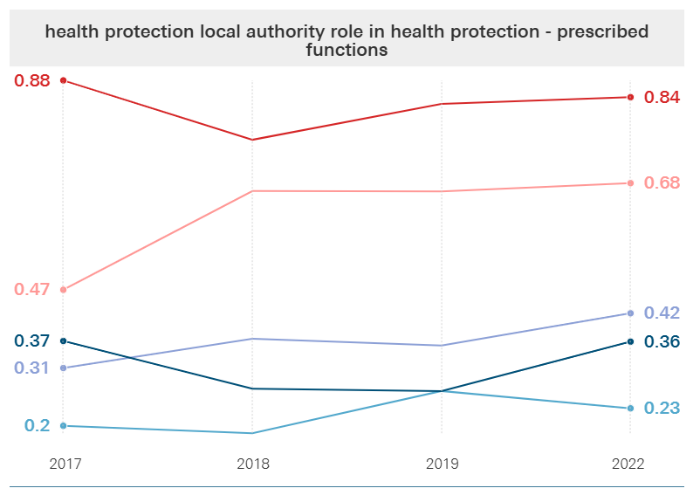 | 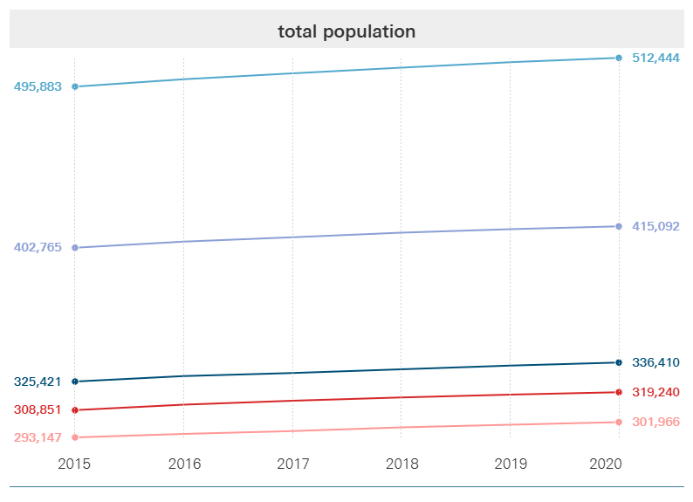 |
| 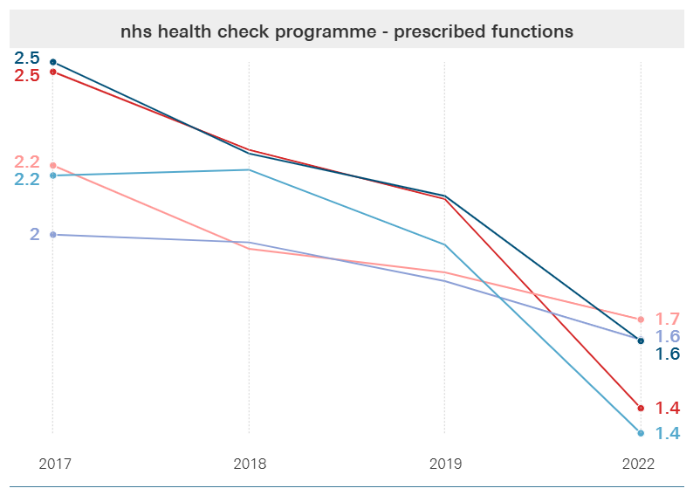 | 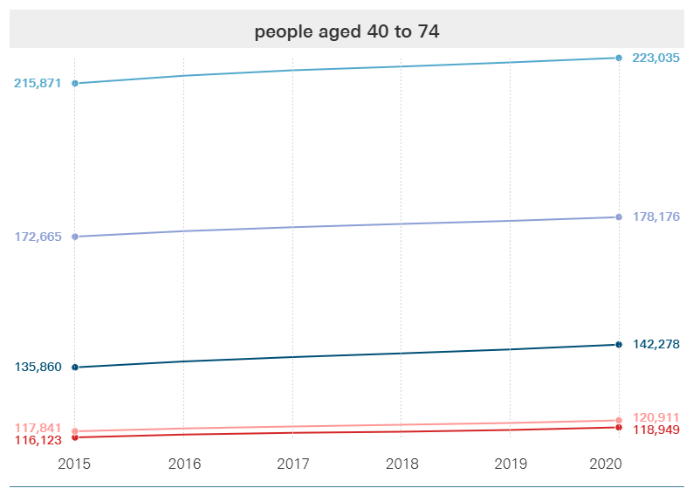 |
| 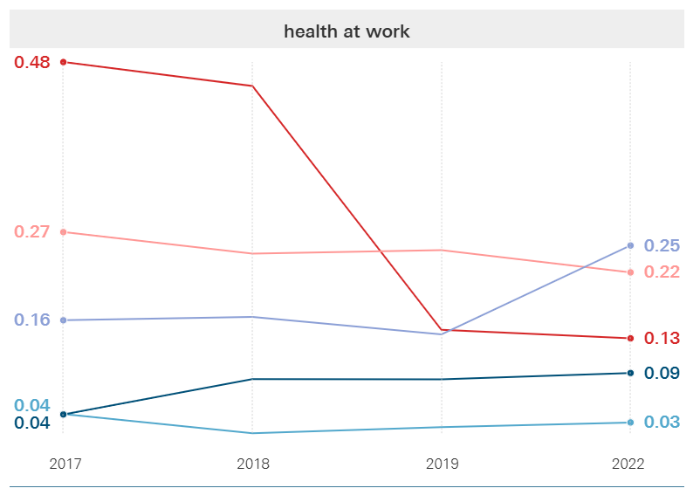 | 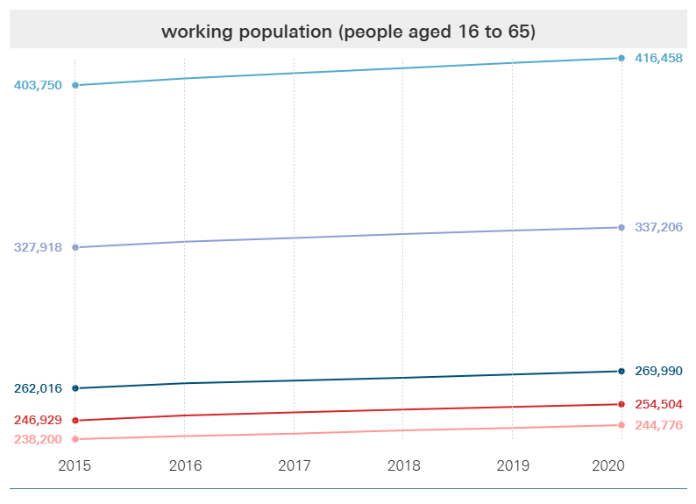 |
| 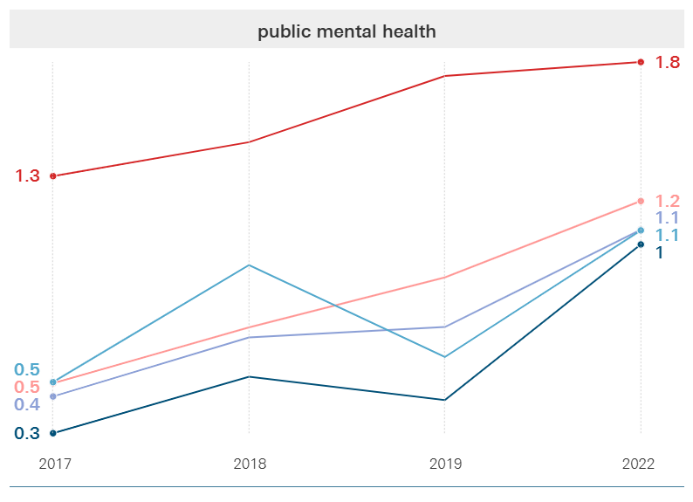 | 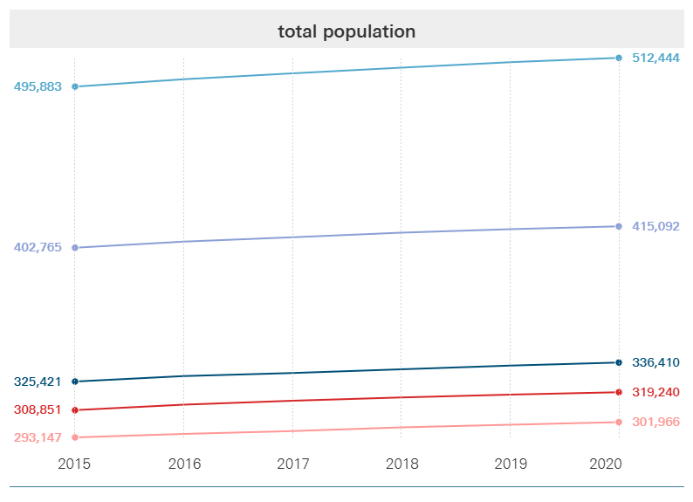 |
| 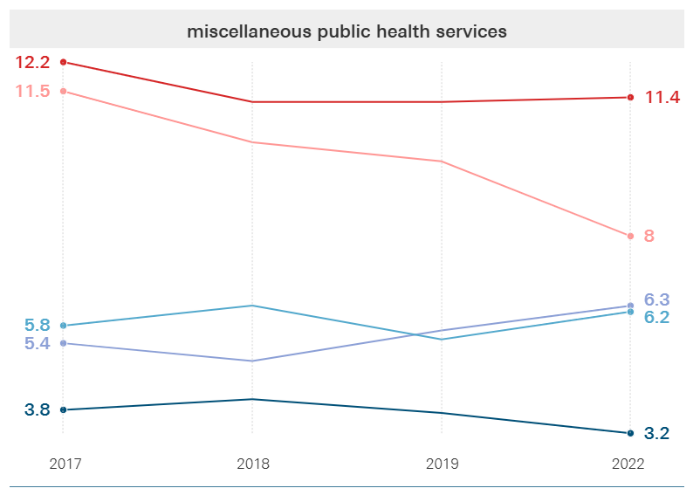 | 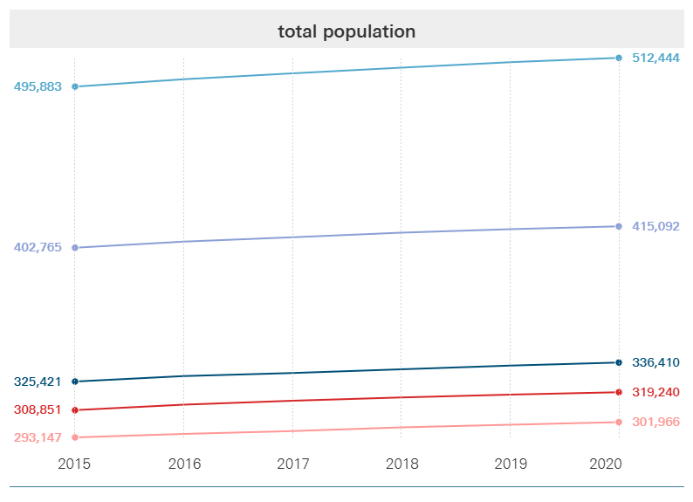 |
| 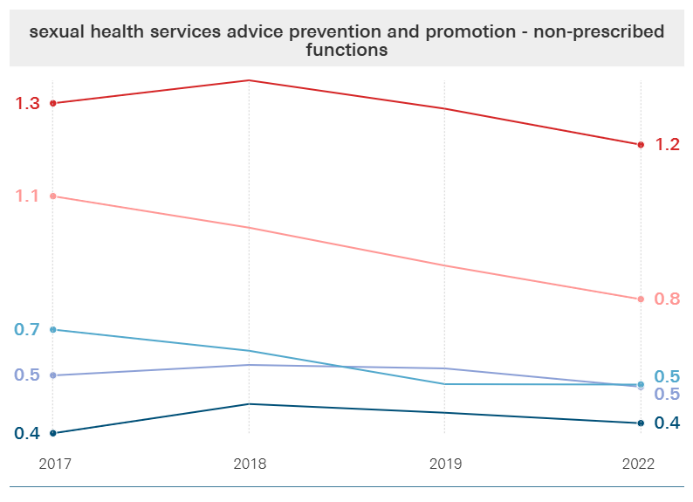 | 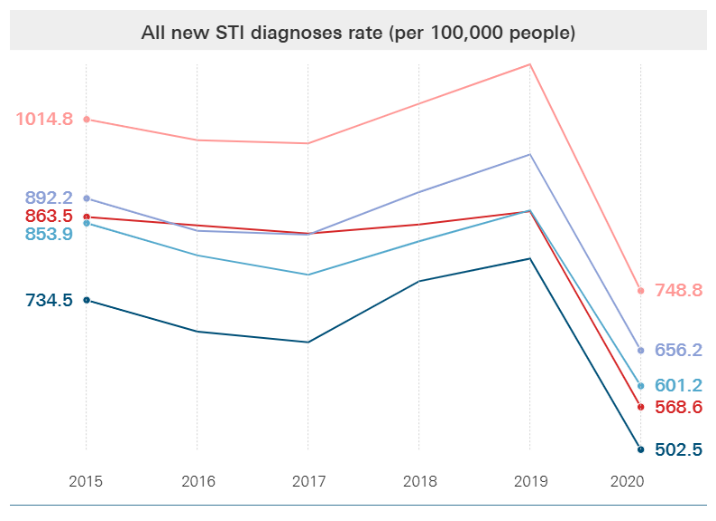 |
| 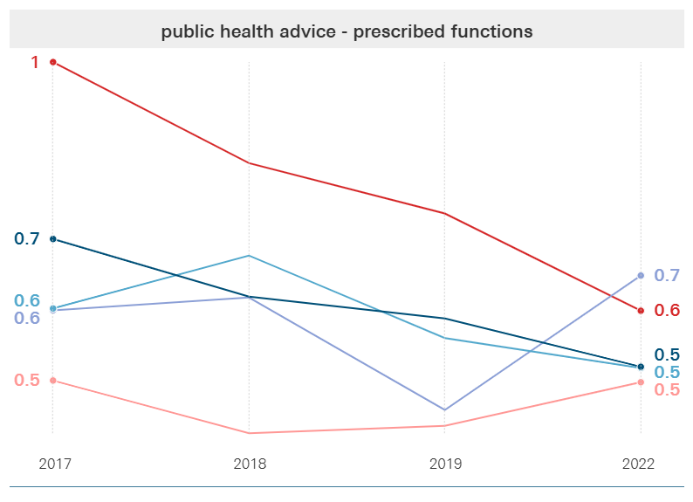 | 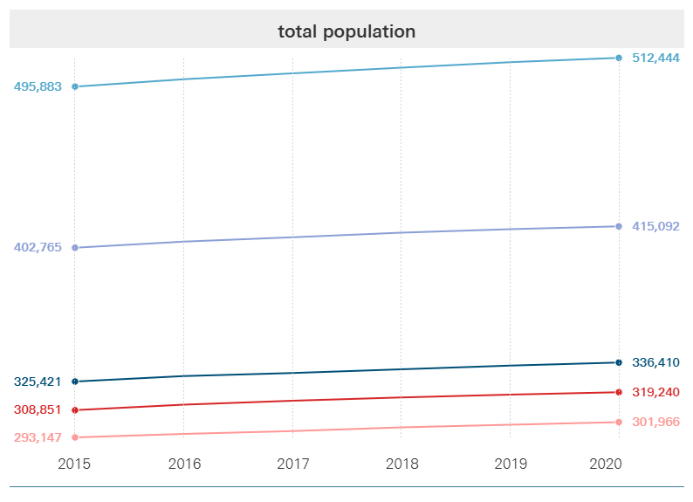 |

**Appendix 4: Sensitivity Analyses**

We performed sensitivity analyses where the lag between indicators of need and spending year was altered to one year and three years. As can be seen below, altering these lags did not result in substantial changes:

| **Variable** | **One year lag** | | **Three year lag** | |
| --- | --- | --- | --- | --- |
|  | Pillai’s Trace | P-value | Pillai’s Trace | P-value |
| Urbanisation category | 0.51 | <0.01*** | 0.40 | <0.01*** |
| Year | 0.97 | <0.01*** | 1.03 | 0.01*** |
| IMD quintile | 0.55 | <0.01*** | 0.50 | <0.01*** |
| Under 18s conception rate per 1,000 | 0.08 | 0.01*** | 0.08 | 0.03*** |
| Year 6 prevalence of obesity | 0.09 | <0.01*** | 0.08 | <0.01*** |
| Alcohol specific mortality | 0.06 | 0.09 | 0.11 | 0.02*** |
| STI diagnoses rate per 100,000 | 0.10 | <0.01*** | 0.08 | 0.04*** |
| Smoking prevalence in adults | 0.06 | 0.17 | 0.04 | 0.54 |
| Obesity prevalence in adults | 0.07 | 0.07 | 0.08 | 0.02*** |
| Deaths from drug misuse | 0.12 | <0.01*** | 0.08 | <0.02*** |
| Admissions for alcohol specific conditions (<18 yrs) | 0.04 | 0.57 | 0.04 | 0.65 |
| Adult population percentage | 0.14 | <0.01*** | 0.10 | <0.01*** |
| Age 0-5 population percentage | 0.15 | <0.01*** | 0.11 | <0.01*** |
| Working age population percentage | 0.12 | <0.01*** | 0.08 | <0.03*** |
| Age 40-74 population percentage | 0.19 | <0.01*** | 0.17 | <0.01*** |
| Age 5-19 population percentage | 0.18 | <0.01*** | 0.12 | <0.01*** |
| Age 4-5 or 10-11 population percentage | 0.06 | 0.09 | 0.06 | 0.24 |
| Age over 65 population percentage | 0.19 | <0.01*** | 0.20 | <0.01*** |

R-squared measures for the 1 year and 3 year lagged models were small, similar to that of the core analysis (0.24 and 0.25 respectively, compared to 0.23).

**Appendix 5: Sub-group Analyses**

For the breakdown of public health activities included in each of the below sub-categorisations, please see the Methods section.

For all sub-categories, urbanisation category, year and IMD quintile were found to have a significant influence on the composition of the grant at the sub-group level. Beyond this, the range of indicators with a significant effect on the composition of the grant for each sub-group were:

- *Children and young people-related spend:* Year 6 prevalence of obesity and obesity prevalence in adults had a significant association with the proportion of the grant spent on services relating to children and young people compared to all other aspects of spend. The proportion of variance explained by the model for these aspects of spend remained small, with 21% of variation explained (R^2^ = 0.21).
- *Substance misuse and tobacco control spend*: Deaths from drug misuse, proportion of the population over the age of 65, the proportion of the population aged 0-5, the proportion of the population aged 5-19, and proportion of the population aged 40-74 were also found to be significantly associated with the proportion of spend going towards activities in this sub-category. The proportion of variance for this aspect of spend explained by the model was highest of all sub-categories by a considerable margin, with almost half of variance explained (R^2^ = 0.47).
- *Sexual health spend*: Deaths from drug misuse, Year 6 prevalence of obesity, conception rate for people under the age of 18, the proportion of the population aged 0-5, proportion of the population over the age of 65, and proportion of the population age 40-74 had significant associations with the proportion of spend on sexual health services. STI diagnoses per 100,000 did not show a significant association. Despite this the proportion of variance explained by the model was higher than for the overall model, with 29% of variance explained (R^2^ = 0.29).
- *Obesity-related spend*: Year 6 prevalence of obesity, STI diagnoses per 100,000, deaths from drugs misuse, proportion of the population over the age of 65 and the proportion of the population aged 0-5 were significantly associated with the proportion of spend on obesity and physical activity services. No significant association was found with prevalence of obesity in adults. The proportion of variance explained by the model was smallest of all sub-categories (R^2^ = 0.19).
- *Other spending*: Almost all indicators were found to have a significant association with this sub-category of spend (aside from under 18s conception rate, obesity prevalence in adults, deaths from drug misuse, admission episodes for alcohol-specific conditions, and proportion of the population aged between 4-5 or 10-11). The proportion of variance explained by the model similar to the overall model (R^2^ = 0.23).
